# Tau-Connectome Subtypes and Solanezumab Response in Preclinical Alzheimer’s Disease

**DOI:** 10.1101/2025.09.13.25335690

**Authors:** Hamid Abuwarda, Wen-Xiang Tsai, Shengxian Ding, Mohamed Elhassan, Xilin Shen, Yize Zhao, R. Todd Constable, Carolyn A. Fredericks

**Affiliations:** Department of Neurology, School of Medicine, Yale University, New Haven, CT, USA; Interdepartmental Neuroscience Program, School of Medicine, Yale University, New Haven, CT, USA; Institute of Brain Science, National Yang Ming Chiao Tung University, Taipei, Taiwan; Department of Biostatistics, School of Public Health, Yale University, New Haven, CT, USA; Department of Radiology, School of Medicine, Yale University, New Haven, CT, USA; Department of Biomedical Engineering, School of Medicine, Yale University, New Haven, CT, USA

## Abstract

**Importance:** This study identifies clinically meaningful subtypes within preclinical Alzheimer’s disease (AD) using the functional connectome and introduces tau-predictive functional connectivity as a new tool to stratify risk and treatment response.

**Objective:** To determine whether fMRI–derived functional connectivity can identify subgroups with differential cognitive outcomes and treatment responses among amyloid-positive, cognitively unimpaired adults.

**Design, Setting and Participants:** A post-hoc analysis of the Anti-Amyloid in Asymptomatic Alzheimer’s disease (A4) study, a randomized, double-blinded, placebo-controlled clinical trial. The A4 trial was a multi-center clinical trial in the United States, Canada, Australia, and Japan. This trial enrolled cognitively unimpaired adults aged 65-85, of whom 1,490 had successful functional magnetic resonance imaging data preprocessing: A*β* -(n = 445) and A*β* + (n = 1045). Longitudinal data were drawn from both the primary trial and the open-label time period (up to 455 weeks). Hierarchical clustering was performed on baseline functional connectivity patterns, followed by spline-based generalized linear modeling to evaluate differences in longitudinal clinical and biomarker outcomes.

**Intervention(s) or Exposure(s):** Tau-predictive functional connectivity, derived from previously generated connectome-based tau predictive models, was used to cluster individuals at baseline. Subsequent analyses compared longitudinal outcomes within the placebo-only group and within the treatment group.

**Main Outcome(s) and Measure(s):** For the original A4 trial, the primary endpoint was the Preclinical Alzheimer’s Cognitive Composite (PACC) score, with secondary clinical outcomes including the Clinical Dementia Rating Sum of Boxes (CDR-SB), Cognitive Function Index (CFI), and Activities of Daily Living (ADL). Secondary biomarker endpoints included plasma phosphorylated tau-217 (pTau-217) and cortical amyloid SUVR (Standardized Uptake Value Ratio). We assessed significance using either permutation testing or bootstrapped models (n=5000). All confidence intervals were defined by bootstrapped models.

**Results:** Hierarchical clustering of connectivity data revealed two distinct subgroups among amyloid-positive individuals in the A4 trial. Individuals had distinct tau PET spatial distributions: one subgroup (Cluster 1, n=618) with a typical limbic-predominant distribution, and the other (Cluster 2, n=427) showing an atypical cortical-predominant distribution. In the placebo arm, the cortical-predominant group exhibited significantly more cognitive decline on both PACC (marginal mean difference: -3.5, 95% CI: -6.4 to -0.6, p = 0.009) and secondary cognitive and clinical outcomes (CDR-Sum of Boxes: +1.8 (+0.6 to +3.1), p =0.005; Activities of Daily Living: - 8.7 (-16.5 to -1.2), p = 0.010; Cognitive Function Index: +4.4 (+1.1 to +7.9) p=0.010; Plasma pTau-217 not significant: p = 0.67; FDR-adjusted for 4 tests) at the end of the study (Week 455). Remarkably, in the cortical-predominant cluster, those treated with solanezumab had significantly improved PACC scores compared to placebo counterparts (marginal mean difference at 455 weeks: +3.3, 95% CI: +0.4 to +6.4, p = 0.03). This corresponds to 48% less cognitive decline, equivalent to a 103-week difference in modeled cognitive trajectories over the 9-year study period, despite the trial’s overall null results. Secondary clinical outcomes were not significant in either cluster. In typical tau group, solanezumab showed no effect in secondary outcomes (CDR-SB: -0.1, 95% CI: –0.8 to +0.7; p=0.80; ADL: +0.9, 95% CI: –3.3 to +5.1 p=0.80; CFI: –0.2, 95% CI: –2.1 to +1.5, p=0.80; FDR-adjusted for 5 tests). In the cortical-predominant group, effects trended in favor of solanezumab on CDR-SB: -1.1, 95% CI: –0.4 to +2.8; p=0.20; ADL: +2.5, 95% CI: –6.4 to +12.1; p=0.60; CFI: –2.5, 95% CI: –6.4 to +0.8; p=0.20; FDR-adjusted for 5 tests) but none reached significance. Amyloid clearance was significantly reduced in both groups at equal levels (Typical tau Solanezumab-Placebo *Δ*SUVR: -0.06, 95% CI: -0.08 to -0.04, p =<0.001; Cortical-predominant tau Solanezumab-Placebo *Δ*SUVR: -0.05, 95% CI: -0.08 to -0.02, p = 0.008; FDR-adjusted for 5 tests). There was a significant reduction in plasma pTau-217 for cortical-predominant solanezumab patients (-0.29; 95% CI: -0.49 to -0.11, p=0.008; FDR-adjusted for 5 tests), while the typical group showed no associated differences (-0.08; 95% CI: -0.22 to +0.05, p = 0.63; FDR-adjusted for 5 tests).

**Conclusions and Relevance:** We show that baseline clustering using tau-predictive functional connectivity identified an AD subtype with worse overall outcomes and stronger responsiveness to solanezumab. These findings highlight the potential of the functional connectome to personalize interventions in AD and in future clinical trials.

**Key Points:** *Question:* Can tau-predictive functional connectome patterns derived from baseline functional magnetic resonance imaging (fMRI) identify clinically meaningful subgroups in preclinical Alzheimer’s disease?

*Findings:* We performed a post-hoc analysis of the Anti-Amyloid in Asymptomatic Alzheimer’s (A4) study, a clinical trial of solanezumab in preclinical Alzheimer’s patients. Hierarchical clustering of baseline tau-predictive functional connectome signatures identified two distinct subgroups. Groups showed no baseline differences in demographic or biomarker variables, except for a modest difference in racial distribution. Spatial tau distributions differed among the groups, with one group showing a classical limbic-predominant pattern and the other having a cortical-predominant tau pattern. Among participants in the placebo arm, the cortical-predominant subgroup demonstrated poorer longitudinal scores on the A4 trial’s primary endpoint, the Preclinical Alzheimer’s Cognitive Composite (-3.3 points; lower score indicates greater cognitive impairment), relative to the other group. Importantly, within the treatment arm, individuals belonging to the cortical predominant subgroup exhibited positive treatment effects relative to the placebo group, despite the trial’s overall null results.

*Meaning:* Our clustering analyses used the baseline fMRI-derived functional connectome to identify a vulnerable subgroup in preclinical AD individuals that benefited from the drug in the A4 null trial of solanezumab. These findings support the use of fMRI-based biomarkers for patient stratification and the development of more personalized treatments in future clinical trials of Alzheimer’s disease.

## Introduction

Preclinical Alzheimer’s disease (AD) is a clinically asymptomatic state that can last decades, offering a critical window for intervention to preserve neural circuitry and cognition. Recent disease-modifying therapeutics, specifically anti-amyloid therapies like lecanemab and donanemab, have ushered in a new era of AD treatments^1,2^. However, early efforts to intervene during the preclinical stage, such as the A4 trial, have yielded null results^3^. Encouragingly, lecanemab and donanemab, which target more mature amyloid species, are currently under investigation in preclinical AD^4,5^. Taken together, studies to date underscore both the promise and challenges of interventions during this stage.

Techniques that can identify groups of individuals most likely to benefit from therapies are critical for personalizing treatments. For example, a recent trial of blarcamesine demonstrated greater clinical benefit in patients with wildtype sigma-1 receptor compared to the overall trial population^6^. This illustrates how stratification on biologically relevant features can reveal treatment effects that may be obscured in the full cohort. Past work has used multiple modalities, such as fluid biomarker assays, PET, structural MRI, neuropsychiatric metrics, and genetics, in a data-driven manner to identify latent subgroups in clinical and preclinical disease; while informative, their clinical effectiveness has not yet been established^7–10^. Functional magnetic resonance imaging (fMRI), which defines an individual’s functional connectome^11^, is underutilized to identify clinically meaningful AD subgroups.

In this study, we aimed to address that gap. We previously showed that functional connectome-based models predict focal tau deposition in preclinical AD and generalize to the Alzheimer’s Disease Network Initiative-3 cohort^12^. We hypothesized that these tau-predictive connectivity features are meaningful to disease progression and could define clinically meaningful subgroups in preclinical AD. Using these predictive edges, we applied unsupervised clustering to baseline fMRI data of amyloid-positive participants in the A4 trial and assessed longitudinal outcomes and treatment response in the resulting subgroups.

This data-driven approach identified two distinct subtypes, which did not differ on key demographics including APOE status, baseline cortical amyloid load, or baseline plasma biomarkers (including pTau-217). One subtype showed tau PET patterns consistent with the typical limbic-predominant distribution (Cluster 1), while the other had an atypical cortical-predominant distribution (Cluster 2). Evaluation of longitudinal outcomes on placebo showed that this cortical-predominant subtype exhibited worse cognitive and clinical outcomes over time relative to the typical group. Remarkably, patients in the cortical-predominant group on solanezumab had higher longitudinal PACC scores compared to placebo counterparts, a trend not seen in treatment arms of the typical tau group. This work highlights tau-predictive functional connectivity as a promising tool for stratifying preclinical AD patients who may differ in both disease trajectory and therapeutic responsiveness in future clinical trials.

## Methods

### Datasets and Participants

We conducted a post-hoc analysis of data from the Anti-Amyloid in Asymptomatic Alzheimer’s disease (A4) study^3^. The A4 study enrolled amyloid-PET positive, cognitively to evaluate unimpaired adults aged 65-85 years the efficacy of solanezumab, an anti-monomeric amyloid antibody. Of 1,648 baseline participants with resting-state fMRI, 1,490 passed QA (1045 amyloid-positive, 445 amyloid-negative). Amyloid-negative participants were used for normative modeling and to generate reference curves; clustering analyses were performed exclusively on the amyloid-positive group. Site information was unavailable.

### Hierarchical Clustering

We adapted a hierarchical clustering method (Figure 1) from Tozzi et al.^13^, using a repeated 5-fold cross-validation approach to evaluate model/parameter combinations. Candidate solutions were assessed with adjusted Rand indices (stability) and silhouette scores (cohesion/separability). We identified the optimal solution using these metrics to define consensus connectivity features. Using these features, we calculated individual deviations of normative patterns, which were used as inputs to the clustering algorithm.

**Figure 1.**
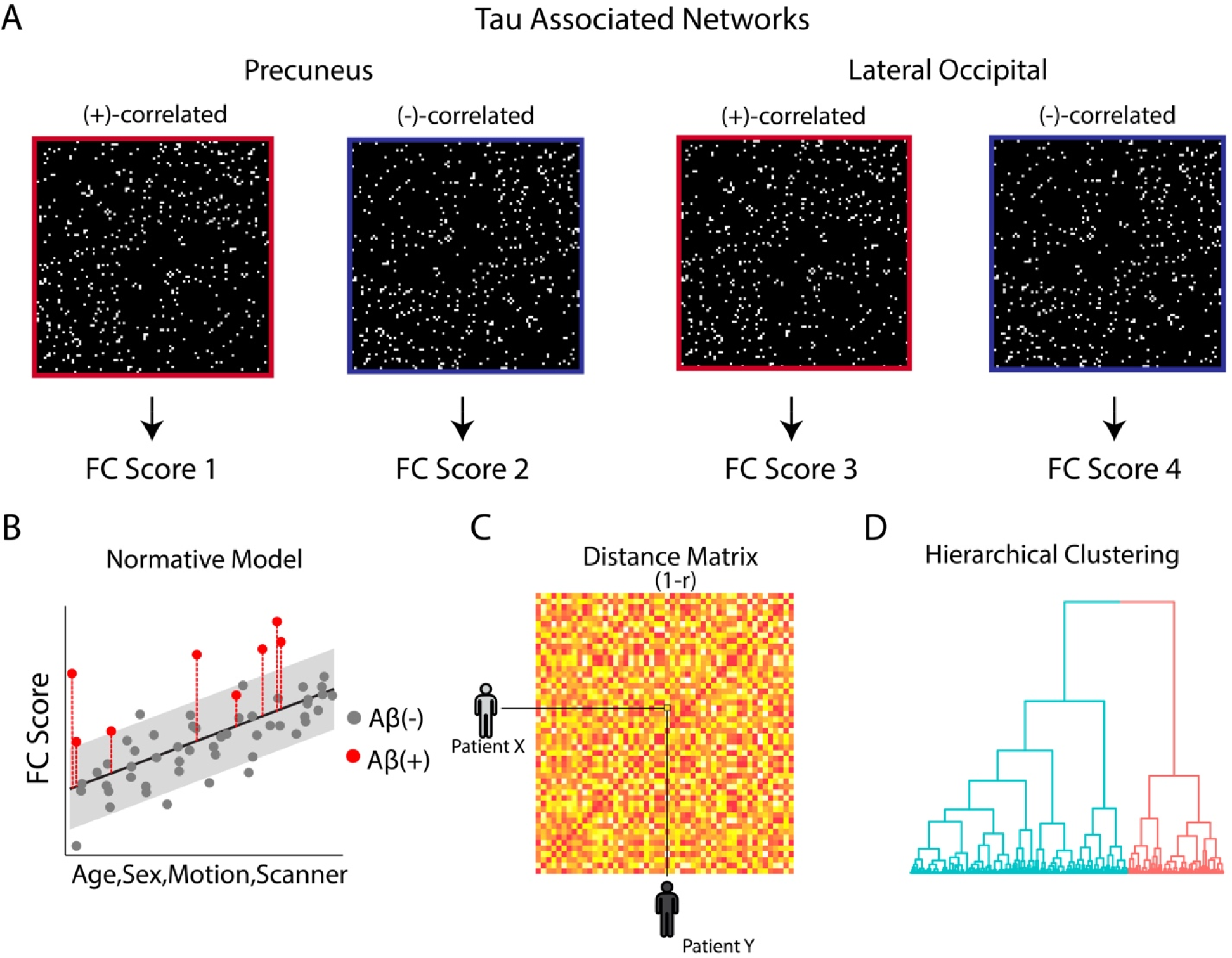
Approach schematic. A) Two connectome-based predictive models feature sets, split into positively and negatively correlated edges to regional tau, were used. Connectivity values from each group of edges were averaged to generate four connectivity scores. B) Connectivity scores were modeled as a function of age, sex, motion, and scanner in the amyloid-negative individuals, representing expected brain connectivity patterns of these connectivity scores for these covariates in a healthy control population. A*β*+ patients were then applied to this model, and Z-scores for each of the four connectivity scores for each patient were generated. C) Distance matrix was computed, summarizing the dissimilarity of deviation scores between participants in the amyloid-positive group. D) Hierarchical clustering was applied to the distance matrix, revealing natural groupings of subjects based on their connectivity profiles.

### Model Selection

Previously, we generated connectome-based predictive models^14^ of focal tau PET^12^. We tested 14 total models of focal tau, of which six demonstrated significance against permutation testing: posterior cingulate, precuneus, lateral occipital, middle temporal, inferior temporal, and bank of the superior temporal sulcus. In the present analysis, we systematically evaluated all possible combinations of the six connectome-based models of regional tau z-scores to identify the most stable, separable and cohesive clustering solution. Because Pearson correlation was used to define the distance matrix (see ’Clustering’), at least two regional models were required. A single model yields two datapoints per subject, producing trivial correlations of +/-1. Including a minimum of two models (and therefore four datapoints) ensured sufficient variability for stable distance estimation.

We chose cluster solutions that simultaneously optimized stability, cohesion, and separability as indicators of valid subgroups^15^. To assess stability, we used the Adjusted Rand Index (ARI), a measure of agreement between clustering solutions that corrects for chance, ranging from 1 (perfect agreement), and -1 (worse than chance)^16^. To assess cohesion and separability, we used silhouette scores, which quantify how similar each subject is to its assigned cluster (optimizing cohesion) compared to other clusters (optimizing separation), ranging from -1 (misclassified) to 1 (well classified)^17^.

We applied a leave-20%-out cross-validation approach to evaluate all model combinations across thresholds for edge selection in the tau-connectome predictive models (5-fold cross-validation, repeated 1000 times). Cross-validation thresholds of 3 or 4 out of 5, and iteration thresholds of 600, 700, 800, and 900 out of 1000 were tested. Clusters with an ARI >0.65 when compared to the full cohort clustering results (suggesting moderate cluster recovery/stability) were retained. Of these, we selected the model combination that generated the most cohesive and separable clusters, defined by silhouette score. The model combination yielding the highest silhouette score incorporated predictive features from the precuneus and lateral occipital models, with edges thresholded at 3 out of 5 cross-validation folds and 600 out of 1000 iterations.

### Feature Extraction

Using these optimal models and connectivity thresholds, we generated consensus masks for edges positively and negatively correlated with precuneus and lateral occipital tau, as these sets showed distinct spatial distributions (e.g., negatively correlated tau predictive edges more commonly identified in nodes of higher-order association areas) (Figure 1B)^12^. Mean connectivity values within each edge set was modeled as a function of age, sex, head motion, and scanner using a regression model built in the amyloid-negative LEARN cohort (n = 445), serving as a normative reference. For each amyloid-positive patient, we calculated deviation from this healthy control normative model (scaled by the LEARN residual standard deviation), generating four functional connectivity z-scores per participant (Figure 1B).

### Clustering

We computed a correlation-based distance matrix based on these z-scores, where the distance between two participants was defined as *d* = 1 - *r*, with *r* representing the Pearson’s correlation coefficient (Figure 1C). Agglomerative hierarchical clustering, a bottom-up approach iteratively merging clusters with the smallest average pairwise distance, with average linkage was applied (Figure 1D)^13^. We evaluated solutions ranging from two to five clusters and selected the optimal number based on silhouette scores. We chose two clusters, as 96% of all model combinations evaluated found that the two-cluster solution was optimal.

### Longitudinal Analyses

To test the relationship between clusters and longitudinal outcomes, we assessed trajectories of the A4 study’s primary and secondary endpoints, comparing individuals in Cluster 1 and Cluster 2 in both the placebo (to assess for differences in outcome by cluster with no drug) and drug (to assess for differences in drug effect by cluster) groups. The Preclinical Alzheimer’s Cognitive Composite (PACC) was the primary endpoint of the A4 trial. Secondary cognitive and clinical endpoints included the Clinical Dementia Rating Sum of Boxes (CDR-SB), the Cognitive Function Index (CFI), and Activities of Daily Living (ADL). Biomarker outcomes were assessed using plasma pTau-217 and amyloid-beta PET. Outcomes were modeled using natural cubic splines, similar to the original A4 study^3^, with minor modifications (Supplemental Methods). Tau heat maps for PACC longitudinal analyses of treatment effects were generated by including a precuneus tau z-score interaction term (instead of subgroup label).

### Statistical Analysis

Statistical significance for group differences was assessed using permutation tests to test for group exchangeability. Cluster assignments were, randomly while preserving the original cluster sizes, iterated 5000 times to generate a null distribution, and the marginal mean difference at Week 455 was assessed against this distribution. Bootstrapping (n=5000 iterations) was used to assess stability of contrast estimates, generate confidence intervals, and assess significance between treatment arms within each cluster. PACC was not corrected for the false-discovery rate (FDR) as it was the primary endpoint of the study. Analyses for secondary endpoints were FDR-corrected using the Benjamini-Hochberg procedure. Details are described in Supplemental Methods. Amyloid PET significance was assessed by two-sided t-test of groupwise difference, and confidence intervals were estimated by the corresponding 95% confidence limits.

## Results

Hierarchical clustering of tau-predictive functional connectivity features from the A4 trial yielded two stable, cohesive, and separable groups (mean silhouette score = 0.72, mean ARI = 0.83). The two-cluster solution was the most common across all cross-validation runs, which was optimal in 96% of model combination evaluations as measured by silhouette scores. Of the 1045 amyloid-positive patients, 68% (n = 618) were assigned to Cluster 1 and 32% (n = 427) to Cluster 2.

The two clusters did not differ in baseline demographics or key AD risk markers, including age, sex, APOE 𝛜4 carrier status, baseline amyloid burden, baseline biomarkers (plasma pTau-217, GFAP, NFL, and amyloid-42/40), or baseline PACC (Table 1), as assessed using two-sample t-tests for continuous variables and chi-square or Fisher’s exact tests, as appropriate, for categorical variables. Race distributions differed between clusters and were therefore included as a covariate in all subsequent analyses.

**Table 1.** Participant Baseline Characteristics.

|  | (n=445)<br>Amyloid-negative | (n=618)<br>Amyloid-Positive<br>Cluster 1 | (n=427)<br>Amyloid-Positive<br>Cluster 2 | Cluster 1 vs Cluster<br>2 FDR-adjusted p-value |
| --- | --- | --- | --- | --- |
| **Age, yrs (sd)** | 70.51 (4.41) | 71.81 (4.75) | 72.33(4.97) | 0.36 |
| **Female, no.**<br>**(%)** | 278 (62.5%) | 376 (60.8%) | 243(56.9%) | 0.44 |
| **Education, yrs**<br>**(sd)** | 16.71 (2.58) | 16.65 (2.71) | 16.45(2.88) | 0.46 |
| **Race, no. (%)** |  |  |  | <0.001 |
| White | 416 (93.5%) | 591 (95.6%) | 386(90.4%) |  |
| Black | 10 (2.2%) | 16 (2.6%) | 12(2.8%) |  |
| Asian | 10 (2.2%) | 3 (0.5%) | 24(5.6%) |  |
| Other | 9 (2.0%) | 8 (1.3%) | 5(1.2%) |  |
| **Ethnic Group,**<br>**no. (%)** |  |  |  | 0.41 |
| Hispanic or<br>Latino | 17 (3.8%) | 18 (2.9%) | 16(3.7%) |  |
| Not Hispanic or<br>Latino | 427 (96.0%) | 596 (96.4%) | 403(94.4%) |  |
| Unknown or not<br>reported | 1 (0.2%) | 4 (0.6%) | 8(1.9%) |  |
| **PACC, (sd)** | 0.78 (2.35) | 0.16(2.63) | -0.19(2.71) | 0.30 |
| **APOE** |  |  |  | 0.92 |
| **Genotype, no.**<br>**(%)** |  |  |  |  |
| $\epsilon 2/\epsilon 3$ | 50(11.2%) | 30(4.9%) | 25(5.9%) | |
| $\epsilon 2/\epsilon 4$ | 9(2.0%) | 22(3.6%) | 16(3.7%) | |
| $\epsilon 3/\epsilon 3$ | 286(64.3%) | 216(35.0%) | 155(36.3%) | |
| $\epsilon 3/\epsilon 4$ | 92(20.7%) | 297(48.1%) | 192(45.0%) | |
| $\epsilon 4/\epsilon 4$ | 3(0.7%) | 52(8.4%) | 38(8.9%) | |
| $\epsilon 2/\epsilon 2$ | 5(1.1%) | 1(0.2%) | 1(0.2%) | |
| **Amyloid**<br>**Centiloid, (sd)** | 4.52(12.52) | 65.34(33.39) | 66.87(31.61) | 0.68 |
| **Plasma GFAP,**<br>**(sd)** | 0.10(0.05) | 0.12(0.06) | 0.13(0.06) | 0.41 |
| **Plasma NF-κL,**<br>**(sd)** | 3.13(1.36) | 3.36(1.66) | 3.40(1.47) | 0.85 |
| **Plasma**<br>**Aβ 42/40, (sd)** | 105.39(20.28) | 91.55(18.30) | 90.81(14.06) | 0.76 |
| **Plasma**<br>**ptau 217, (sd)** | NaN (NA) | 0.29(0.15) | 0.30(0.16) | 0.73 |

Average tau PET spatial distributions differed across clusters, in which one (Cluster 1) showed a spatial pattern resembling the typical limbic-predominant pattern (Figure 2A). By contrast, Cluster 2 showed a cortical-predominant phenotype, with especially high tau levels in nodes such as the precuneus and posterior cingulate cortex (Figure 2A). We henceforth refer to Cluster 1 as the ‘typical’ tau group and Cluster 2 as the ‘cortical-predominant’ tau group.

**Figure 2.**
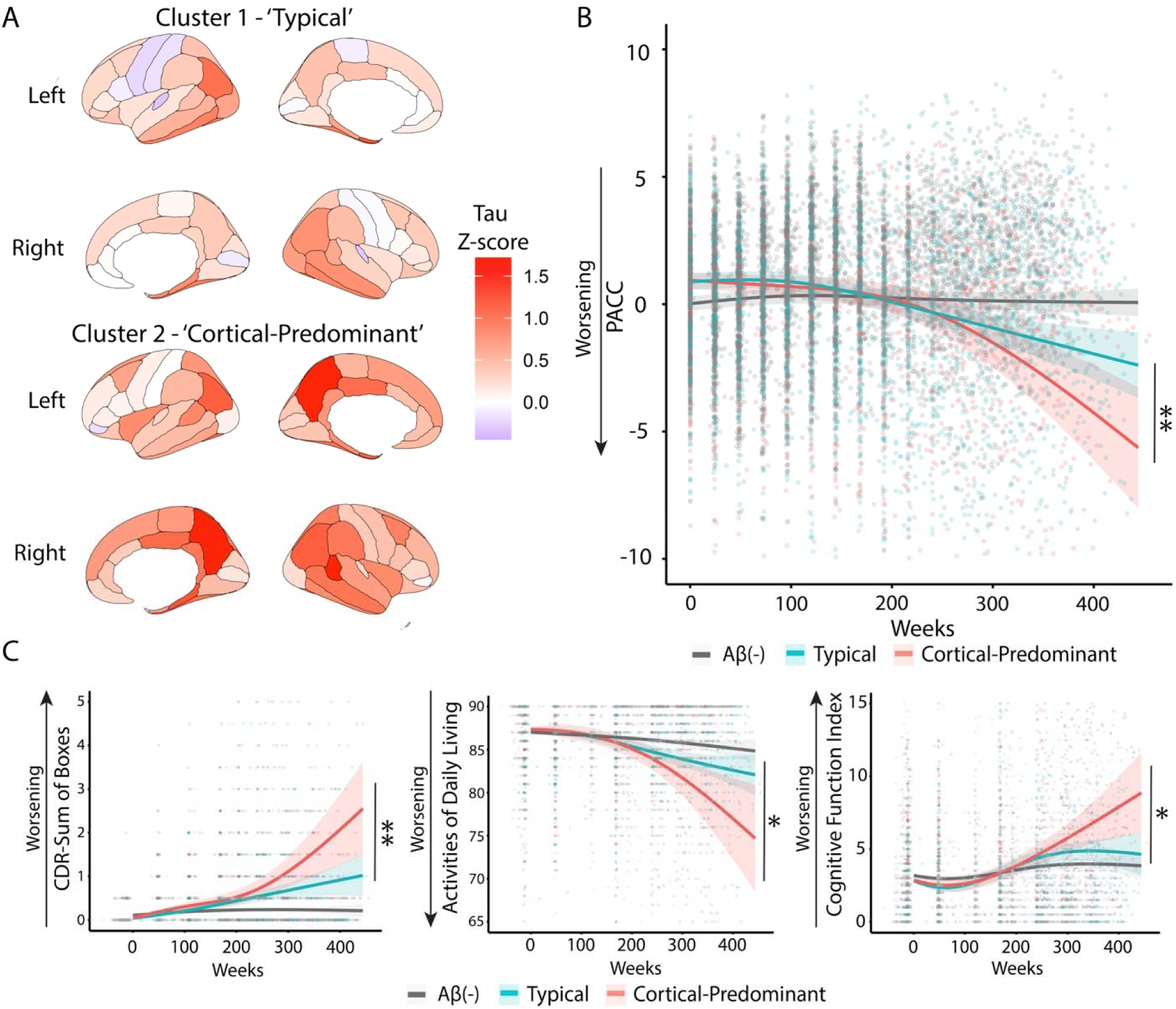
Placebo outcomes by cluster A) Tau patterns by subtype, calculated as the average z-score tau PET signal (relative to amyloid-negative participants) across all individuals classified as each subtype. B) Trajectories of the preclinical Alzheimer’s Cognitive Composite (PACC) over 9 years in typical tau (blue) and cortical-predominant (red) subgroups. Each point represents an individual participant score, with shaded bands showing 95% confidence intervals. B) Secondary outcomes by clusters, including Clinical Dementia Rating-Sum of Boxes (CDR-SB), Activities of Daily Living (ADL), and Cognitive Function Index (CFI). Shaded bands represent 95% confidence intervals. * = p<0.05, ** = p<0.01 by permutation testing.

We next evaluated outcome differences between these groups. We first assessed cluster differences in the placebo arm to assess natural disease trajectories without treatment influence. At the study end (Week 455), cortical-predominant tau groups scored on average 3.3 points lower on the Preclinical Alzheimer’s Cognitive Composite (PACC) than typical participants (95% CI: -0.8 to -6.0 p<0.001) (Figure 2B).

The cortical-predominant subgroup also demonstrated greater functional and subjective decline on secondary outcome tests compared to the typical tau group (CDR-SB: +1.8 points, 95% CI: +0.6 to +3.1, p=0.005; ADL: -8.7 points, 95% CI: -16.5 to -1.2, p=0.010; CFI: +4.4 points, 95% CI: +1.1 to +7.9; p=0.010; FDR-adjusted for 4 permutation tests). These between-group differences at the study end were not accompanied by differences in longitudinal plasma pTau-217 trajectories (Table 2; -0.03; 95% CI: -0.14 to 0.09; p = 0.67; FDR-adjusted for four permutation tests).

**Table 2.** Summary of Longitudinal Outcomes.

|  | Estimated Difference (95% CI) | p-value |
| --- | --- | --- |
| **Placebo: Cortical – Typical** |  |  |
| PACC | -3.5 (-6.4, -0.6) | 0.009 |
| *Secondary Outcomes* |  |  |
| CDR-SB* | +1.8 (+0.6, +3.1) | 0.005 |
| ADL * | -8.7 (-16.5, -1.2) | 0.01 |
| CFI* | +4.4 (+1.1, +7.9) | 0.01 |
| Plasma pTau-217* | -0.03 (-0.14, +0.09) | 0.67 |
| **Typical: Solanezumab – Placebo** |  |  |
| PACC | +0.1 (-1.7, +2.1) | 0.90 |
| *Secondary Outcomes* |  |  |
| CDR-SB* | -0.1 (-0.8, 0.7) | 0.78 |
| ADL * | +0.9 (-3.3, +5.1) | 0.68 |
| CFI* | -0.2 (-2.1, +1.5) | 0.80 |
| Plasma pTau-217* | -0.08 (-0.22, +0.05) | 0.25 |
| Relative Amyloid Reduction* | -0.058 (-0.079, -0.037) | <0.001 |
| **Cortical-Predominant: Solanezumab – Placebo** |  |  |
| PACC | +3.3 (+0.4, +6.4) | 0.03 |
| *Secondary Outcomes* |  |  |
| CDR-SB* | -1.1 (-0.4, +2.8) | 0.16 |
| ADL * | +2.5 (-6.4, +12.1) | 0.60 |
| CFI* | -2.5 (-6.4, +0.8) | 0.15 |
| Plasma pTau-217* | -0.29 (-0.49, -0.11) | <0.001 |
| Relative Amyloid Reduction* | -0.045 (-0.075, -0.015) | 0.003 |

\* = FDR-adjusted

We next assessed whether treatment responses differed by cluster membership for the primary and secondary endpoints. For the primary cognitive endpoint (PACC), solanezumab was associated with attenuated PACC decline in cortical-predominant group (Figure 3A; marginal mean difference at week 455: +3.3; 95% CI: +0.4 to +6.4; p = 0.03). This corresponded to a 48% decrease in cognitive decline and cognitive-time saved of 103 weeks relative to placebo over the course of the 455-week study period. The time corresponding to a 30% reduction in cognition, the original A4 study PACC endpoint, was reached in the cortical-predominant group at 280 weeks (about 5.4 years). The typical tau group showed no associated treatment effect (Figure 3A; week 455 marginal mean difference: +0.01; 95% CI: -1.7 to +2.1; p = 0.90). Assessing PACC scores by precuneus tau load in the tau PET cohort highlighted that the associated treatment differences were greatest in those with the highest cortical tau levels (Figure 3B).

**Figure 3.**
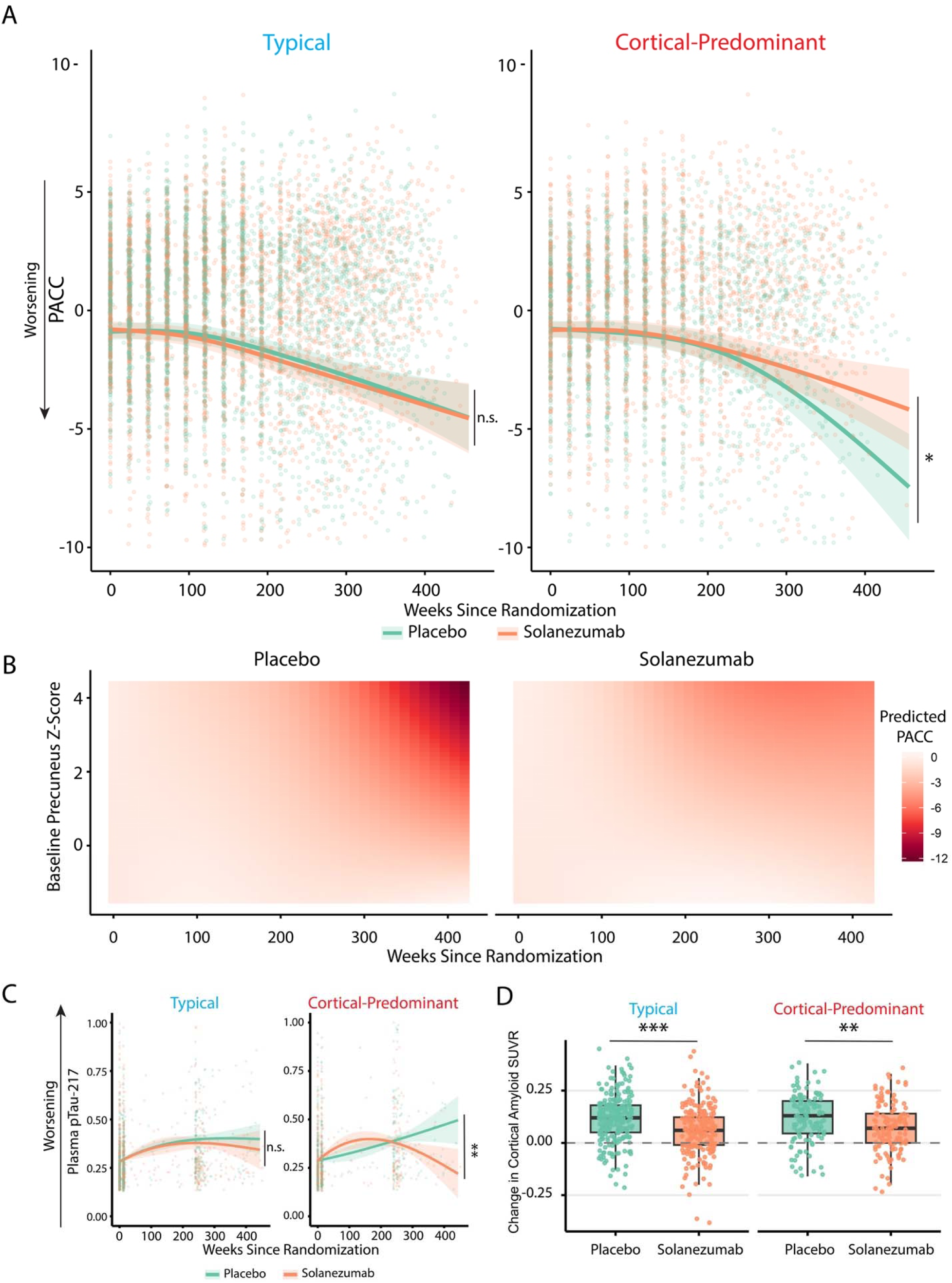
Longitudinal outcomes of subtypes by treatment arm. A) Change in PACC over time is shown, with trajectories plotted separately for solanezumab (orange) and placebo (teal). Individual data points are overlaid with fitted lines and 95% confidence intervals. B) Heatmaps of modeled PACC scores over time by precuneus tau z-scores. C) Plasma phosphotau-217 trajectories plotted separately for solanezumab and placebo groups. Individual data points are overlaid with fitted lines and 95% confidence intervals. D) Change in cortical amyloid SUVR levels in placebo and solanezumab groups by subgroup (week 240 – baseline). Boxes denote interquartile ranges, and dots represent individual participants. * = p<0.05, ** = p<0.01, *** = p<0.001

The cortical-predominant solanezumab participants also had significantly lower pTau-217 compared to placebo (Figure 3C; -0.29; 95% CI: -0.49 to -0.11, p=0.008; FDR-adjusted for 5 bootstrap tests). The typical tau group showed no effect (Figure 3C, -0.08; 95% CI: -0.22 to +0.05, p = 0.63; FDR-adjusted for 5 bootstrap tests). Secondary clinical measures showed patterns favoring solanezumab treatment in the cortical-predominant group, though these did not reach statistical significance (Table 1). Both clusters demonstrated equivalent amyloid mitigation with solanezumab (Figure 3D; Typical: -0.06, 95% CI: -0.08 to -0.04, p<0.001; Cortical-Predominant: -0.05, 95% CI: -0.08 to -0.02, p = 0.008; FDR-adjusted for 5 bootstrap tests).

## Discussion

Anti-amyloid antibodies have emerged as the first disease-modifying treatments for Alzheimer’s disease, with therapies such as lecanemab and donanemab showing benefits in early symptomatic stages^1,2^. Ongoing trials are now testing whether these drugs, which target amyloid aggregates, are also effective in preclinical AD patients^4,5^. Previously, the A4 trial failed to show that monomeric amyloid removal during the preclinical phase provided cognitive benefits. Our findings, however, suggest that treatment benefits may be confined to clinically meaningful subgroups in preclinical disease.

We provide converging lines of evidence of clinically meaningful subgroups in preclinical AD. First, we use data-driven clustering on tau-predictive functional connectivity features to identify two subgroups, with no differences in baseline age, sex, cognition, plasma tau, or global amyloid burden, but with different tau distributions: one showing a typical tau distribution, while the other has more cortical-predominant tau, particularly in the precuneus. The two clusters demonstrate differences in naturalistic longitudinal outcomes, with the cortical-predominant group performing worse. Most importantly, over nearly nine years of follow-up, participants in the cortical-predominant tau group appear to benefit from solanezumab, with an associated 48% reduction in cognitive decline and approximately 2 years of cognitive time-saved relative to their placebo counterparts; the limbic group, consistent with the original study, showed no meaningful benefit.

There are several important implications of this study. We show that fMRI-based clustering methods may have practical utility in identifying individuals more likely to benefit from therapies. MRI is widely available and already integrated into clinical workflows for patients receiving anti-amyloid therapies. Adding a baseline functional sequence could enable stratification and enrich trials for individuals most likely to respond, thus increasing power and salvaging therapeutic effects. While a prior post-hoc analysis of the A4 trial examined treatment effects in relation to global tau PET signal with no benefit, our approach has identified data-driven clusters that captured interactive effects between tau burden and treatment response likely obscured by global measures alone over longer follow-up^18^. The cortical-predominant group aligns with cortical-predominant/hippocampal-sparing subtypes previously described^19–21^ and may be one of two fundamental disease progression pathways in addition to a limbic-predominant pathway^22^, mirroring the two fundamental subtypes we identify in this work. The cortical-predominant phenotype has been associated with more aggressive disease, including more rapid cognitive decline and earlier death^19,21^, corresponding with our findings of poorer longitudinal outcomes.

Current models propose that tau pathology preferentially propagates along functionally connected pathways, particularly as it extends out of the medial temporal lobe into parietal hubs such as the precuneus^23,24^. The cortical-predominant subtype may represent a network state especially vulnerable to rapid, connectivity-mediated tau spread. In this context, solanezumab, hypothesized to act as a peripheral sink of monomeric amyloid, may shift aggregation dynamics to reduce oligomeric amyloid, which has been shown to be synaptotoxic and facilitate synaptic tau uptake^25–28^. Consequently, individuals more vulnerable to connectivity-mediated tau propagation may experience disproportionate benefit, as lowering oligomeric amyloid could attenuate trans-synaptic propagation of tau more effectively than those with slower tau propagation which remains confined to limbic. The fact that end-of-study plasma pTau-217, which correlates with both amyloid and tau PET, was reduced only in the cortical-predominant tau solanezumab patients, supports the presence of an underlying biological treatment effect in this subgroup.

### Limitations

In this post-hoc analysis, randomization was not stratified by cluster, so confounding factors cannot be fully excluded. While a similar number of placebo and solanezumab patients within each cluster argues against imbalance, a prospective trial is needed to validate findings. The cognitive benefit observed in cortical-predominant patients on solanezumab emerged at around 5.5 years, which may reflect the long temporal window of preclinical AD. Therefore, in preclinical cohorts, the identification of responsive subgroups in preclinical AD could help power clinical trials by reducing heterogeneity. Improvements in fMRI acquisition, such as longer scan durations and inclusion of task-based paradigms, can also enhance connectivity estimates and cluster assignment accuracy^29,30^; 30 minutes of resting-state fMRI scan time has recently been supported as optimally balancing power and scan cost and is worth considering for future studies^31^. While pTau-217 was significantly different between subgroups at end-of-study, the sparsity of tau data points compared to cognitive measures limits the precision of this estimate. Additional biomarker sampling in future trials can confirm this observation and allow better discrimination of relationship between early tau and connectivity changes. At the time of publication, the A4 study was the only large preclinical dataset with longitudinal clinical, biomarker, and fMRI data. We previously generalized our connectome-based tau predictive models to the ADNI-3 cohort^12^, but no such alternate large preclinical dataset exists for the validation of the treatment effects we observe here. As additional preclinical trial datasets become available, external validation will be critical, especially in more cognitively and demographically diverse samples that better represent the clinical population. Post-hoc analyses of lecanemab did not show benefits in early symptomatic AD when stratifying by cortical tau burden^32^, suggesting disease stage or targeted amyloid species are important considerations. Future work can help determine whether this improvement in this cortical subtype outcomes extends to other disease-modifying therapies in preclinical disease.

## Conclusions

Despite the negative results of the preclinical A4 study, we demonstrate cognitive benefit for solanezumab in one of two subgroups defined using tau-predictive functional connectivity. The subgroup with a cortical-predominant tau distribution (32% of cohort) had significantly reduced cognitive decline on solanezumab, showing a 48% reduction in cognitive decline and 2-year difference in modeled cognitive decline over 9 years compared with placebo; the other subgroup showed no benefit, consistent with the overall results of the trial. These results suggest that the functional connectome, which is easily derived from a widely available 6-minute MRI sequence, and its relationship to early tau can identify preclinical AD patients at-risk for increased cognitive decline and those who may specifically benefit from treatments. Incorporating functional connectomics into trial design could reduce heterogeneity and improve power by enabling the identification of clinically meaningful subgroups for targeted intervention.

## Data Availability

Data is available through the A4 website

https://www.a4studydata.org/

## Notes

### Competing Interest Statement

The authors have declared no competing interest.

### Funding Statement

HHS | NIH | National Institute of General Medical Sciences (NIGMS), HHS | NIH | National Institute of General Medical Sciences (NIGMS), HHS | NIH | National Institute of General Medical Sciences (NIGMS), HHS | NIH | National Institute of General Medical Sciences (NIGMS), HHS | NIH | National Institute of General Medical Sciences (NIGMS), HHS | NIH | National Institute of General Medical Sciences (NIGMS), HHS | NIH | National Institute of General Medical Sciences (NIGMS)

### Author Declarations

A4 data was openly available dataset

## References

1. Sims JR, Zimmer JA, Evans CD, et al. Donanemab in Early Symptomatic Alzheimer Disease: The TRAILBLAZER-ALZ 2 Randomized Clinical Trial. JAMA. 2023;330(6):512–527. doi:10.1001/jama.2023.13239

2. Dyck CH van, Swanson CJ, Aisen P, et al. Lecanemab in Early Alzheimer’s Disease. New England Journal of Medicine. Published online January 5, 2023. doi:10.1056/NEJMoa2212948

3. Sperling RA, Donohue MC, Raman R, et al. Trial of Solanezumab in Preclinical Alzheimer’s Disease. New England Journal of Medicine. 2023;389(12):1096–1107. doi:10.1056/NEJMoa2305032

4. Khartabil N, Awaness A. Targeting Amyloid Pathology in Early Alzheimer’s: The Promise of Donanemab-Azbt. Pharmacy (Basel). 2025;13(1):23. doi:10.3390/pharmacy13010023

5. Rafii MS, Sperling RA, Donohue MC, et al. The AHEAD 3-45 Study: Design of a prevention trial for Alzheimer’s disease. doi:10.1002/alz.12748

6. Macfarlane S, Grimmer T, Teo K, et al. Blarcamesine for the treatment of Early Alzheimer’s Disease: Results from the ANAVEX2-73-AD-004 Phase IIB/III trial. The Journal of Prevention of Alzheimer’s Disease. 2025;12(1):100016. doi:10.1016/j.tjpad.2024.100016

7. Alashwal H, El Halaby M, Crouse JJ, Abdalla A, Moustafa AA. The Application of Unsupervised Clustering Methods to Alzheimer’s Disease. Front Comput Neurosci. 2019;13. doi:10.3389/fncom.2019.00031

8. Ferreira D, Nordberg A, Westman E. Biological subtypes of Alzheimer disease. Neurology. 2020;94(10):436–448. doi:10.1212/WNL.0000000000009058

9. Wisch JK, Butt OH, Gordon BA, et al. Proteomic clusters underlie heterogeneity in preclinical Alzheimer’s disease progression. Brain. 2023;146(7):2944–2956. doi:10.1093/brain/awac484

10. Petersen KK, Nallapu BT, Lipton RB, Grober E, Ezzati A. MRI-guided clustering of patients with mild dementia due to Alzheimer’s disease using self-organizing maps. NeuroImage: Reports. 2024;4(4):100227. doi:10.1016/j.ynirp.2024.100227

11. Finn ES, Shen X, Scheinost D, et al. Functional connectome fingerprinting: identifying individuals using patterns of brain connectivity. Nat Neurosci. 2015;18(11):1664–1671. doi:10.1038/nn.4135

12. Abuwarda H, Trainer A, Horien C, et al. Whole-brain functional connectivity predicts regional tau PET in preclinical Alzheimer’s disease. Brain Commun. 2025;7(4):fcaf274. doi:10.1093/braincomms/fcaf274

13. Tozzi L, Zhang X, Pines A, et al. Personalized brain circuit scores identify clinically distinct biotypes in depression and anxiety. Nat Med. 2024;30(7):2076–2087. doi:10.1038/s41591-024-03057-9

14. Shen X, Finn ES, Scheinost D, et al. Using connectome-based predictive modeling to predict individual behavior from brain connectivity. Nat Protoc. 2017;12(3):506–518. doi:10.1038/nprot.2016.178

15. Hennig C. Cluster validation by measurement of clustering characteristics relevant to the user. *arXiv*. Preprint posted online September 8, 2020. doi:10.48550/arXiv.1703.09282

16. Hubert L, Arabie P. Comparing partitions. Journal of Classification. 1985;2(1):193–218. doi:10.1007/BF01908075

17. Rousseeuw PJ. Silhouettes: A graphical aid to the interpretation and validation of cluster analysis. Journal of Computational and Applied Mathematics. 1987;20:53–65. doi:10.1016/0377-0427(87)90125-7

18. Sperling RA, Donohue MC, Rissman RA, et al. Amyloid and Tau Prediction of Cognitive and Functional Decline in Unimpaired Older Individuals: Longitudinal Data from the A4 and LEARN Studies. The Journal of Prevention of Alzheimer’s Disease. 2024;11(4):802–813. doi:10.14283/jpad.2024.122

19. Young CB, Winer JR, Younes K, et al. Divergent Cortical Tau Positron Emission Tomography Patterns Among Patients With Preclinical Alzheimer Disease. JAMA Neurology. 2022;79(6):592–603. doi:10.1001/jamaneurol.2022.0676

20. Shand C, Markiewicz PJ, Cash DM, et al. Heterogeneity in Preclinical Alzheimer’s Disease Trial Cohort Identified by Image-based Data-Driven Disease Progression Modelling. medRxiv. Preprint posted online February 10, 2023:2023.02.07.23285572. doi:10.1101/2023.02.07.23285572

21. Murray ME, Graff-Radford NR, Ross OA, Petersen RC, Duara R, Dickson DW. Neuropathologically defined subtypes of Alzheimer’s disease with distinct clinical characteristics: a retrospective study. The Lancet Neurology. 2011;10(9):785–796. doi:10.1016/S1474-4422(11)70156-9

22. Poulakis K, Pereira JB, Muehlboeck JS, et al. Multi-cohort and longitudinal Bayesian clustering study of stage and subtype in Alzheimer’s disease. Nat Commun. 2022;13(1):4566. doi:10.1038/s41467-022-32202-6

23. Vogel JW, Iturria-Medina Y, Strandberg OT, et al. Spread of pathological tau proteins through communicating neurons in human Alzheimer’s disease. Nat Commun. 2020;11(1):2612. doi:10.1038/s41467-020-15701-2

24. Roemer-Cassiano SN, Wagner F, Evangelista L, et al. Amyloid-associated hyperconnectivity drives tau spread across connected brain regions in Alzheimer’s disease. Science Translational Medicine. 2025;17(782):eadp2564. doi:10.1126/scitranslmed.adp2564

25. Lacor PN, Buniel MC, Chang L, et al. Synaptic Targeting by Alzheimer’s-Related Amyloid β Oligomers. J Neurosci. 2004;24(45):10191–10200. doi:10.1523/JNEUROSCI.3432-04.2004

26. Shankar GM, Bloodgood BL, Townsend M, Walsh DM, Selkoe DJ, Sabatini BL. Natural Oligomers of the Alzheimer Amyloid-β Protein Induce Reversible Synapse Loss by Modulating an NMDA-Type Glutamate Receptor-Dependent Signaling Pathway. J Neurosci. 2007;27(11):2866–2875. doi:10.1523/JNEUROSCI.4970-06.2007

27. Shankar GM, Li S, Mehta TH, et al. Amyloid-β protein dimers isolated directly from Alzheimer’s brains impair synaptic plasticity and memory. Nat Med. 2008;14(8):837–842. doi:10.1038/nm1782

28. Kadamangudi S, Marcatti M, Zhang WR, et al. Amyloid-β oligomers increase the binding and internalization of tau oligomers in human synapses. Acta Neuropathol. 2024;149(1):2. doi:10.1007/s00401-024-02839-2

29. Noble S, Scheinost D, Finn ES, et al. Multisite reliability of MR-based functional connectivity. NeuroImage. 2017;146:959–970. doi:10.1016/j.neuroimage.2016.10.020

30. Greene AS, Gao S, Scheinost D, Constable RT. Task-induced brain state manipulation improves prediction of individual traits. Nat Commun. 2018;9(1):2807. doi:10.1038/s41467-018-04920-3

31. Ooi LQR, Orban C, Zhang S, et al. Longer scans boost prediction and cut costs in brain-wide association studies. Nature. 2025;644(8077):731-740. doi:10.1038/s41586-025-09250-1

32. Charil A, Cao Y, Willis BA, Hersch S, Irizarry MC, Reyderman L. Lecanemab Slows Tau PET Accumulation. Alzheimer’s & Dementia. 2024;20(S6):e091956. doi:10.1002/alz.091956

