## Supplemental for "Tau-Connectome Subtypes and Solanezumab Response in Preclinical Alzheimer’s Disease"

**Supplemental Methods**

*MRI Acquisition and Preprocessing* Imaging was performed on 3T MRI scanners, and the acquisition included resting-state fMRI (echo-planar imaging sequence, 6.5 minutes duration) and T1-weighted anatomical scans. Complete scanning parameters and protocols are available through the A4 study data website: https://www.a4studydata.org/.

Image preprocessing followed established pipelines detailed previously (Abuwarda *et al.,* 2025, Ju *et al.,* 2023). Skull stripping of anatomical images was performed using optiBET, after which images were nonlinearly registered to MNI-152 space. Functional images with available slice acquisition parameters underwent slice time correction. All images were motion corrected, linearly registered to anatomical space, and spatially smoothed. We only retained scans with mean frame-to-frame displacement less than 0.3 mm to minimize artifacts. Nuisance signal regression removed motion parameters, physiological noise, and scanner drift. Finally, functional connectivity matrices were generated using the Shen-268 atlas (Shen *et al.,* 2013).

*Models* The primary and each secondary endpoint measure was modeled with a generalized least squares regression:

$$\text{Endpoint}_{ij}=\beta_{0}+f\left( \text{Time}_{ij} \right)*\text{Group}_{i}+\beta_{C}\text{Covariates}_{i}+\epsilon_{ij},$$

where,

- $\boldsymbol{\beta}_{0}$ represents the intercept
- $f\left( \text{Time}_{ij} \right)*\text{Group}_{i}$ represents the natural cubic splines (df=3) of time with group-specific interactions
- $\boldsymbol{\beta}_{C}\text{Covariates}_{i}$ represents the covariates (age, education, race, APOE-e4 carrier status, baseline cortical amyloid, and baseline outcome score (with test version included for PACC models)
- $\varepsilon_{ij}$ represents the residual error with structured covariance (heteroscedastic by time, continuous auto-regressive (CAR(1)) correlation structure within subjects.

Race was added as an additional covariate given baseline differences between groups. Models were evaluated with 2, 3, and 4 natural cubic splines; 3 was determined optimal by Akaike Information Criterion (AIC). The model failed to converge using either an unstructured or an autoregressive moving average correlation structure, so a CAR(1) structure was implemented. Models were fit by restricted maximum likelihood. Aside from these model differences, the model was the same as the original A4 study (Sperling et al. 2023). Confidence intervals were estimated using robust sandwich estimators with Satterthwaite degrees of freedom. To assess for treatment outcomes, we employed the same models as above but added an additional treatment interaction term, creating a 3-way time, group, and treatment interaction. Raw phosphorylated tau-217 was modeled similarly with the following modifications: natural cubic spline = 2 (optimal by AIC) and covariates included APOE-e4, baseline ptau-217, race, and age.

*Statistics* Permutation significance was assessed with the following:

$$P=\frac{\#\left\{ \left| \hat{\theta}_{\text{null}} \right|\geq\left| \hat{\theta}_{\text{obs}} \right| \right\}}{N}$$

where,

- $\hat{\theta}_{\text{null}}$ represents the null distribution
- $\hat{\theta}_{\text{obs}}$ represents the observed value
- $N$ represents the number of permutations
- $P$ represents the permutation $p$-value

To assess stability of contrast estimates and generate confidence intervals, bootstrap resampling (n=5000 iterations) was performed. Patients in each group were iteratively resampled with replacement while maintaining the within-subject correlation structure. Bootstrap p-values were calculated as twice the minimum proportion of bootstrap estimates on either side of zero, providing a two-tailed test. 95% confidence intervals are defined by bootstrap 2.5th and 97.5th percentiles of the bootstrap distribution.

**Treatment Effect Size and Timing Analyses**. Percent reduction in cognitive decline was calculated as (drug decline - placebo decline)/placebo decline x 100, where cognitive decline represents the change from baseline to Week 455. Cognitive-time difference between the groups was estimated by using the Week 455 marginal mean PACC score in Cluster 2 solanezumab patients, identifying the corresponding time in which Cluster 2 placebo participants achieved this same marginal mean PACC score, and taking the difference between the two timepoints.

**Software**. Clustering analyses were performed in MATLAB (9.11.0.2022996 (R2021b) Update 4). All statistical analyses were performed in R (RStudio Version 2024.12.0+467).

**Acknowledgements**

The A4 Study was a secondary prevention trial in preclinical Alzheimer's disease, aiming to slow cognitive decline associated with brain amyloid accumulation in clinically normal older individuals. The A4 Study was funded by a public-private-philanthropic partnership, including funding from the National Institutes of Health-National Institute on Aging, Eli Lilly and Company, Alzheimer's Association, Accelerating Medicines Partnership, GHR Foundation, an anonymous foundation, and additional private donors, with in-kind support from Avid Radiopharmaceuticals, Cogstate, Albert Einstein College of Medicine and the Foundation for Neurologic Diseases.The companion observational Longitudinal Evaluation of Amyloid Risk and Neurodegeneration (LEARN) Study was funded by the Alzheimer's Association and GHR Foundation. The A4 and LEARN Studies were led by Dr. Reisa Sperling at Brigham and Women's Hospital, Harvard Medical School, and Dr. Paul Aisen at the Alzheimer's Therapeutic Research Institute (ATRI) at the University of Southern California. The A4 and LEARN Studies were coordinated by ATRI at the University of Southern California, and the data are made available under the auspices of Alzheimer’s Clinical Trial Consortium through the Global Research \& Imaging Platform (GRIP). The complete A4 Study Team list is available on: \href{https://www.actcinfo.org/a4-study-team-lists/}{https://www.actcinfo.org/a4-study-team-lists/}. We would like to acknowledge the dedication of the study participants and their study partners who made the A4 and LEARN Studies possible.
